# Clinical performance of automated machine learning: a systematic review

**DOI:** 10.1101/2023.10.26.23297599

**Authors:** Arun James Thirunavukarasu, Kabilan Elangovan, Laura Gutierrez, Refaat Hassan, Yong Li, Ting Fang Tan, Haoran Cheng, Zhen Ling Teo, Gilbert Lim, Daniel Shu Wei Ting

## Abstract

**Introduction:** Automated machine learning (autoML) removes technical and technological barriers to building artificial intelligence models. We aimed to summarise the clinical applications of autoML, assess the capabilities of utilised platforms, evaluate the quality of the evidence trialling autoML, and gauge the performance of autoML platforms relative to conventionally developed models, as well as each other.

**Methods:** This review adhered to a PROSPERO-registered protocol (CRD42022344427). The Cochrane Library, Embase, MEDLINE, and Scopus were searched from inception to 11 July 2022. Two researchers screened abstracts and full texts, extracted data and conducted quality assessment. Disagreement was resolved through discussion and as-required arbitration by a third researcher.

**Results:** In 82 studies, 26 distinct autoML platforms featured. Brain and lung disease were the most common fields of study of 22 specialties. AutoML exhibited variable performance: AUCROC 0.35-1.00, F1-score 0.16-0.99, AUCPR 0.51-1.00. AutoML exhibited the highest AUCROC in 75.6% trials; the highest F1-score in 42.3% trials; and the highest AUCPRC in 83.3% trials. In autoML platform comparisons, AutoPrognosis and Amazon Rekognition performed strongest with unstructured and structured data respectively. Quality of reporting was poor, with a median DECIDE-AI score of 14 of 27.

**Conclusions:** A myriad of autoML platforms have been applied in a variety of clinical contexts. The performance of autoML compares well to bespoke computational and clinical benchmarks. Further work is required to improve the quality of validation studies. AutoML may facilitate a transition to data-centric development, and integration with large language models may enable AI to build itself to fulfil user-defined goals.

## Introduction

In medicine, machine learning (ML) has been applied in a wide variety of contexts ranging from administration to clinical decision support, driven by greater availability of healthcare data and technological development (1–5). Automated machine learning (autoML) enables individuals without extensive computational expertise to access and utilise powerful forms of AI to develop their own models. AutoML thereby enables developers to focus on curating high quality data rather than optimising models manually, facilitating a transition from model-driven to data-driven workflows (6). AutoML has been posited as a means of improving the reproducibility of ML research, and even generating superior model performance relative to conventional ML techniques (7).

AutoML technologies aim to automate some or all of the ML engineering process which otherwise requires advanced data or computer science skills. The first stage is data preparation, involving data integration, transformation, and cleaning. Next is feature selection, where aspects of the data to be utilised in designing the ML model are decided; this may involve data imputation, categorical encoding, and feature splitting (8). Model selection, training, and optimisation are then executed, with model performance evaluation being critical for identification of an optimal solution. AutoML systems use various methods and optimisation techniques to achieve state-of-the-art performance in some or all of the engineering process, such as Bayesian optimisation, random search, grid search, evolutionary based neural architecture selection, and meta-learning (7,9). The optimised model may then be outputted for further work, such as clinical deployment, explainability analysis, or external validation.

AutoML exhibits four major strengths which may support its application in clinical practice and research. Firstly, individual studies have reported comparable performance of autoML to conventionally developed models (10). This raises the possibility of clinical deployment of autoML models and use in pilot studies preceding further model development. Secondly, autoML may improve the reproducibility of ML research by reducing the influence of human technicians who currently engage with an idiosyncratic process of tuning until a satisfactory result is achieved: supporting a transition toward more reproducible data-centric development (6). Thirdly, the reduction in computational experience and hardware conferred by autoML adoption should act as a major democratising force, providing a much larger number of clinicians with access to AI technology (9). Lastly, the time spent on developing models is significantly reduced with autoML, as manual tuning is abolished— this improves efficiency and facilitates an acceleration of exploratory research to establish potential applications of AI (9).

With the myriad of available autoML tools, democratisation of AI beyond those with clinical and computational expertise is feasible, and potential applications are diverse (9,10). However, rigorous validation is necessary to justify deployment. Here, a systematic review was conducted to examine the performance of autoML in clinical settings. We aimed to evaluate the quality of result reporting; describe the specialties and clinical tasks in which autoML has been applied; and compare the performance of autoML platforms with conventionally developed models, as well as each other.

## Methods

The reporting of this study adheres to PRISMA guidance, and the systematic review protocol was prospectively registered on PROSPERO (identifier CRD42022344427) (11,12). The protocol was amended to use a second quality assessment tool (PROBAST) in addition to DECIDE-AI, as described below.

### Data sources and searches

The Cochrane Library, Embase (via OVID), MEDLINE (via OVID), and Scopus were searched from inception up to 11 July 2022, with no initial restrictions on publication status or type. Our search strategy isolated autoML in clinical contexts with the use of Boolean operators, as detailed in Supplementary Material 1. Before screening, duplicates were removed using Zotero version 6.0.14 (Corporation for Digital Scholarship, Vienna, Virginia, US); and Rayyan (11,13).

### Study selection

Abstract screening was conducted in Rayyan by two independent researchers, with a separate third arbitrator with autoML expertise resolving cases of disagreement (13). Full-text screening was similarly conducted by two researchers with a separate arbitrator, in Microsoft Excel for Mac version 16.57 (Microsoft Corporation, Redmond, Washington, US). The explicit, hierarchical criteria for inclusion during abstract and full-text screening are listed below in descending order, with full details provided in Supplementary Material 2:

1. Is written in the English language.
2. Is a peer-reviewed primary research article.
3. Is not a retracted article.
4. Utilises automated machine learning.
5. AutoML is applied in a clinical context.

### Data extraction and quality assessment

For articles satisfying the inclusion criteria, data extraction was conducted by two researchers; with a first clinical researcher’s work verified by a second computational researcher. Quality assessment was conducted by a single researcher, using implicit criteria based on the Developmental and Exploratory Clinical Investigations of DEcision support systems driven by Artificial Intelligence (DECIDE-AI) framework (14). Risk of bias and concerns regarding applicability were assessed similarly by two researchers using the Prediction model Risk Of Bias ASsessment Tool (PROBAST) framework and guidance questions (15).

Other data collected included citation details; autoML platform; processing location (cloud or local); code intensity of the autoML platform; technical features of the autoML platform; clinical task autoML applied towards; medical or surgical specialty defined anatomically where possible; sources of data used to train and test models; training and validation dataset size; dataset format (*i.e.* structured or unstructured); evaluation metrics used to gauge performance; and benchmark figures if presented such as with comparisons to expert clinician or conventional ML performance. Specifically, figures for area under the receiver operator characteristic curve (AUCROC), F1-score, and area under the precision-recall curve (AUCPR) were gathered. If F1-score was not provided but precision (positive predictive value) and recall (sensitivity) were, F1-score was calculated as the harmonic mean of the two metrics. If metrics were not stated in text form but were clearly plotted in graphical form, figures were manually interpolated using WebPlotDigitizer v4.6.0 (Ankit Rohatgi, Pacifica, California, USA). Metrics were excluded if the source or modality of the tested model was unclear (16). Where two researchers disagreed, resolution was achieved through discussion or as-required arbitration by a third researcher.

### Data synthesis and analysis

A narrative synthesis was conducted because meta-analysis was precluded by heterogeneity of datasets, platforms, and use-cases. Quantitative comparisons of autoML models was based on performance metrics (F1-score, AUCROC, AUCPR) to judge the clinical utility of applied autoML (17). AutoML platforms were compared on the same basis where platforms were applied to an identical task with the same data. A statistically significant difference in metrics was defined as featuring non-overlapping 95% confidence intervals. To establish the congruence between studies’ conclusions and their presented data, the discussion and conclusion sections of each study were appraised by a single researcher to identify if autoML was compared to conventional techniques, and if so whether the comparison favoured autoML, conventional techniques, or neither. AutoML platforms were compared in terms of their requirements and capabilities, with researchers contacted to clarify any questions regarding code intensity, processing location, or data structure. Figures were produced with R version 4.1.2 (R Foundation for Statistical Computing, Vienna, Austria) (18–20), and Affinity Designer version 1.10.4 (Pantone LLC, Carlstadt, New Jersey, USA).

## Results

### Record inclusion

Of 2417 records initially identified, 82 were included in the final analysis (Figure 1) (10,16,21–100). In rare cases, research reports referred to autoML or similar terms in the broader context of ‘ML that automates’, despite not utilising autoML technology: these articles were excluded under criterion 4 (101,102). Other borderline cases considered to be outside the scope of this review based on criterion 5 involved uses of autoML in clinical contexts, but without contributing to patient diagnosis, management, or prognosis. These included a surgical video identification and prediction of biological sex from medical images (103,104).

**Figure 1.**
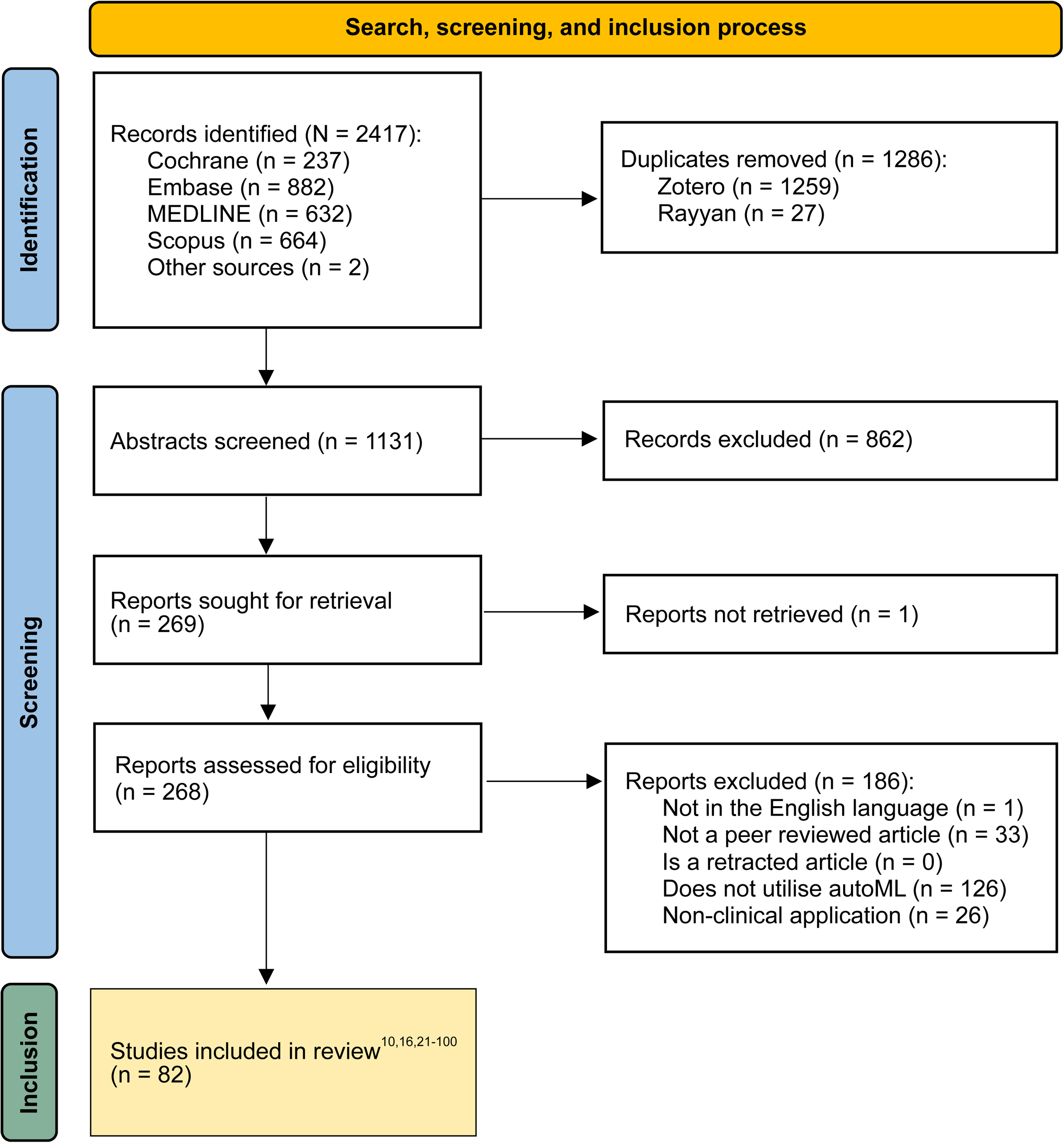
PRISMA flow chart depicting the search, screening, and inclusion process of this review.

### Characteristics of included studies

The characteristics of the 82 included studies are summarised in Figure 2 and Supplementary Material 3. AutoML first entered the medical literature in 2018 and has been growing in impact ever since: 1 paper in 2018; 7 in 2019; 21 in 2020; 35 in 2021; 18 by July 11^th^ 2022. Use-cases are diverse, but diagnostic tasks (53 studies) were more common than management (four studies) or prognostic (25 studies) tasks. The most common specialties in which autoML was used were pulmonology and neurology. Structured (*e.g.* tabulated) and unstructured (*e.g.* imaging) data were used similarly commonly. Dataset size varied widely, between 31 to 2,185,920 for training; 8 to 2,185,920 for internal validation; and 27 to 34,128 for external validation.

**Figure 2.**
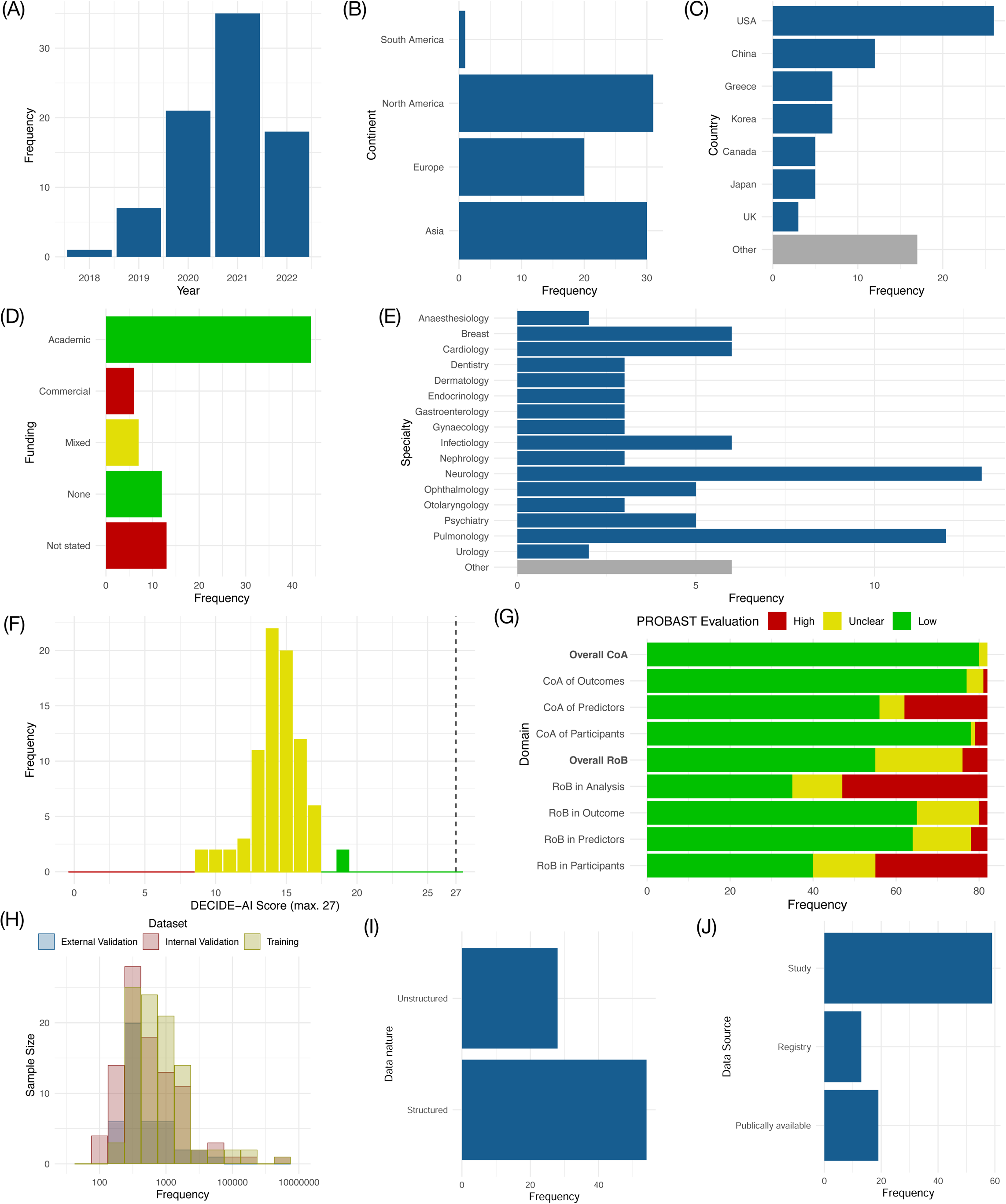
Collectively summarised characteristics of included studies: (A) Date of publication bar chart; (B) Continent of corresponding author bar chart; (C) Country of corresponding author bar chart; (D) Funding source bar chart; (E) Clinical specialty bar chart; (F) DECIDE-AI score histogram; (G) PROBAST evaluation bar chart; (H) Dataset size histogram with logarithmic X-axis; (I) Data nature bar chart; (J) Data source bar chart. RoB = risk of bias; CrA = concerns regarding applicability.

Quality of reporting is summarised in Figure 2F, with individual scores reported in Supplementary Material 4. The median number of fulfilled DECIDE-AI criteria was 14 out of 27, with the highest score being 19 out of 27. Nine criteria were fulfilled by over 90% of included studies. Thirteen criteria were not fulfilled in over half of the included studies: (III) Research governance, (3) Participants, (5) Implementation; (6) Safety and errors in the results; (7) Human factors; (8) Ethics; (VI) Patient involvement; (9) Participants; (10) Implementation; (11) Modifications; (13) Safety and errors in results; (14) Human factors; and (16) Safety and errors in the discussion. Of these, three criteria were not fulfilled by any of the 82 included studies: (8) Ethics; (VI) Patient involvement; and (13) Safety and errors in the results.

Risk of bias and concerns regarding applicability are summarised in Figure 2G. The most common sources of bias were retrospective study design often using publically available datasets, rather than testing autoML models in prospective trials to validate clinical performance and establish generalisability; and failure to provide an appropriate bespoke computational or clinical benchmark to demonstrate the performance of autoML— conferring unclear or high risk of bias in PROBAST appraisal (Supplementary Material 5). In many cases, this was because autoML was used as a tool, rather than the study being a trial of autoML technology, but a statement was made in the discussion or conclusion regarding the effectiveness of autoML in 27 of 47 studies (57%) judged to have a high or unclear risk of bias in the analysis.

### AutoML performance relative to other modalities

The reporting of performance metrics varied widely between papers, likely representing the inherent limitations of applied autoML platforms. 79 studies (96%) provided AUCROC (Figure 3), F1-score (Figure 4), or AUCPR (Supplementary Material 6) as a measure of performance. Of these, 35 studies (44%) exhibited a computational or clinical benchmark to compare autoML performance against, and 21 studies (27%) provided 95% confidence intervals for estimates of performance metrics. Of twelve studies (15%) with benchmark comparisons and confidence intervals, autoML exhibited statistically significantly superior AUCROC in six of 17 trials (35%); significantly superior F1-score in zero of one trial (0%); and significantly superior AUCPR in zero of two trials (0%). In studies with benchmark comparisons and confidence intervals, autoML did not exhibit the lowest AUCROC, F1-score, or AUCPR in any trial. In all studies comparing modalities, autoML exhibited the highest AUCROC in 28 of 37 trials (76%); the highest F1-score in eleven of 26 trials (42%); and the highest AUCPRC in ten of 12 trials (83%). AutoML exhibited the lowest AUCROC in five of 37 trials (14%); the lowest F1-score in six of 26 trials (23%); and the lowest AUCPR in two of twelve trials (17%). For autoML models, AUCROC ranged from 0.346-1.000 (scores of 0.5 are equivalent to chance; maximum score = 1); F1-score ranged from 0.128-0.992 (maximum score = 1); and AUCPR ranged from 0.280-1.000 (maximum score = 1).

**Figure 3.**
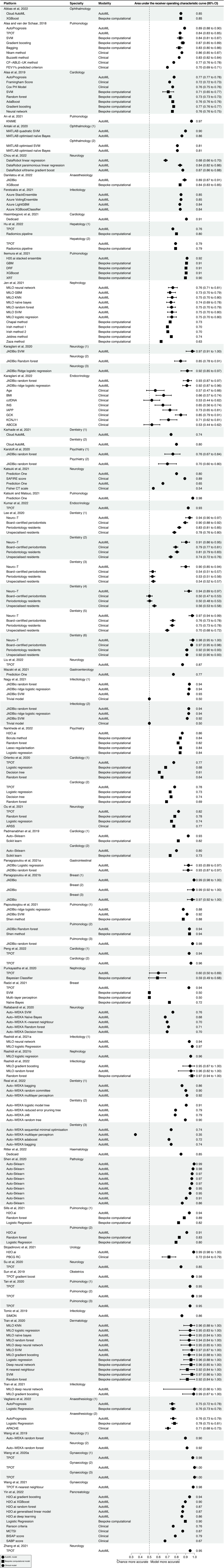
Forest plot depicting reported AUCROC metrics. SVM = support vector machine; FEV1 = forced expiratory volume in 1 second; PH = proportional hazards; GBM = gradient boosting machine; DRF = distributed random forest; XRT = extremely randomised tree; BMI = body-mass index; ccf-DNA = circulating cell-free DNA; CT = computerised tomography; ARSS = Aneurysm Recanalization Stratification Scale; MCTSI = Modified Computed Tomography Severity Index.

**Figure 4.**
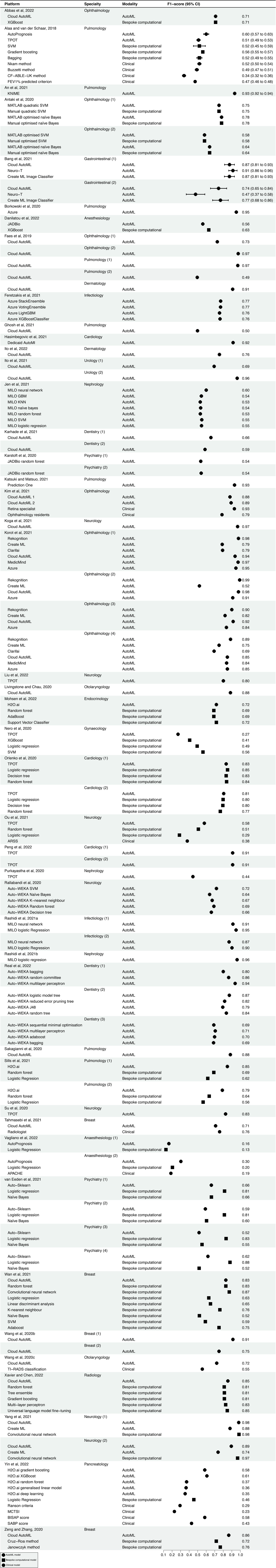
Forest plot depicting reported F1-score metrics. SVM = support vector machine; FEV1 = forced expiratory volume in 1 second; GBM = gradient boosting machine; ARSS = Aneurysm Recanalization Stratification Scale.

57 studies (70%) compared autoML to other conventional modelling methods in the prose of their discussion or conclusion. Of these, 28 suggested that autoML was superior to conventional methods; 29 suggested that autoML was comparable to conventional methods; and none suggested that autoML was inferior to conventional methods. Only 35 studies provided a quantitative comparison in their results, as described above (Figure 3, Figure 4, Supplementary Material 6). Conclusions of comparable effectiveness were justified by congruence with reported performance metrics in 16 of 29 studies (55%); conclusions of superior effectiveness of autoML were justified in eleven of 28 studies (39%).

### Comparative performance of AutoML platforms

A comparative summary of the autoML platforms validated in the literature is presented in Table 1. Platforms vary greatly in their accessibility, technical features, and portability. While performance in different tasks cannot be compared, five studies directly compared distinct autoML platforms in the same task. Of these, one study (20%) provided AUCROC metrics, which favoured AutoPrognosis over TPOT to prognosticate mortality in cystic fibrosis.(23) Four studies (80%) provided F1-score metrics for a total of nine trials (Figure 5): prognosticating mortality in cystic fibrosis; predicting invasion depth of gastric neoplasms from endoscopic photography; diagnosing referrable diabetic retinopathy from fundus photography; diagnosing age-related macular degeneration, central serous retinopathy, macular hole, and diabetic retinopathy from optical coherence tomography (OCT); diagnosing choroidal neovascularisation, diabetic macular oedema, and drusen from OCT; and classifying spine implants from lumbar spine radiographs (23,29,50,97).

**Figure 5.**
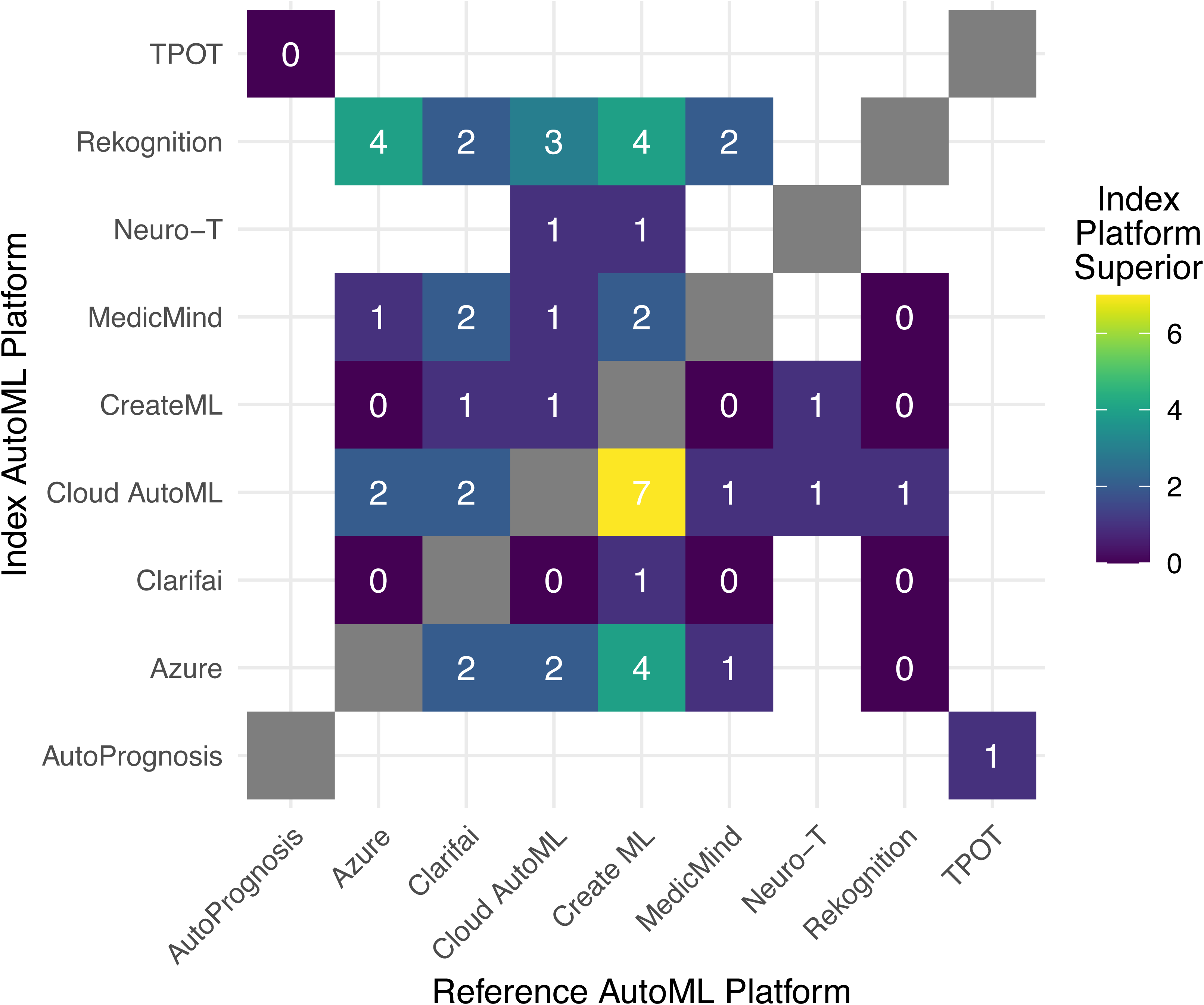
Heat map depicting the comparative performance of autoML platforms as applied to the same clinical tasks in terms of F1-score. Shading and numbers correspond to the number of superior performances exhibited by the index platform with respect to the reference platform.

**Table 1.** Technological comparison of autoML platforms applied in the studies included in this review. AER = approximated error reduction; ML = machine learning; ASSL = automated semi-supervised learning; WEKA = Waikato Environment for Knowledge Analysis; AutoDAL = automated distributed active learning; AutoDC = automated data-centric processing; JADBio = Just-Add-Data Bio; KNIME = Konstanz Information Miner; MILO = Machine Intelligence Learning Optimizer; TPOT = Tree-based Pipeline Optimization Tool; USDM = uncertainty sampling with diversity maximization.

AutoPrognosis (structured data) and Rekognition (unstructured data) exhibited the strongest performance as they were superior to every platform they were compared with, although this was only to TPOT for AutoPrognosis, and Rekognition was compared with fewer platforms than Cloud AutoML. Two studies (40%) reporting five trials provided AUCPR metrics for prognosticating mortality in cystic fibrosis and classifying electrocardiogram traces (23,32). Here, performance favoured AutoPrognosis over TPOT; and AutoDAL-SOAR over USDM, AER, Auto-Weka, Auto-Sklearn, and ASSL+US. While not all platforms can be compared against one another due to incompatibility with data structure, many possible combinations were not trialled and the number of comparative trials was small, making it difficult to establish comparative performance.

### Confidence in conclusions

Confidence in conclusions is tempered by high risk of bias, particularly in retrospective study design and limited metrics facilitating statistical comparisons. However, as autoML did not exhibit statistically significantly worse performance than conventional techniques in any trial and exhibited lower performance metrics than conventional trials in a minority of studies, there is high confidence in the conclusion that autoML technology facilitates production of models with comparable performance to conventional techniques such as bespoke computational approaches. Given the low number of studies providing confidence intervals to enable statistical comparison of models’ performance within trials, conclusions regarding the superiority of autoML relative to conventional techniques have low confidence. In addition, conclusions cannot be assumed to generalise to all use cases and datasets: performance is highly context-specific, as demonstrated by the large variability observed in AUCROC (Figure 3), F1-score (Figure 4), and AUCPR (Supplementary Material 6). Confidence in the superior performance of AutoPrognosis with structured data is very low, as there were very few comparative trials; and low for the superior performance of Rekognition with unstructured data, as the number of comparative trials was low—though not as low as for structured data—and as there were no data for many possible platform comparisons.

## Discussion

This study shows that autoML has been trialled in a wide variety of diagnostic, patient management, and prognostic tasks. AutoML has been used in many clinical specialties, most commonly in brain and lung imaging. Performance of autoML models generally compares well to bespoke computational and clinical benchmarks, often exhibiting superior performance. However, available studies and appraised risk of bias preclude conclusion of autoML providing universally superior performance to conventional modelling; relative and absolute performance vary widely with the applied platform, use case, and data source. The strength of the evidence base supporting use of different autoML platforms is highly heterogenous, with some platforms exhibiting results more supportive of equivalence or superiority to conventional techniques than others. Few studies compared different autoML platforms to determine which provide optimal performance for a given task. Despite these knowledge gaps, a high number of non-comparative studies suggests that autoML is already being applied as a statistical tool, comparable to bespoke machine learning coding packages or statistical software.

There are five main deficiencies in the quality of the autoML evidence base. First, inconsistency in performance metrics may be a consequence of restrictions imposed by autoML platforms but observed variation between studies using similar platforms also suggests that selective reporting is common. Reporting comprehensive metrics is essential, particularly in the context of diagnostic algorithms, as some metrics are a function of prevalence or model threshold (17). Second, explainability analysis is challenging for similar reasons regarding portability, but is possible with emerging technological solutions (22). In addition, some platforms incorporate inbuilt explainability, such as by providing salience maps for DL models. Issues regarding ‘black box’ algorithms are accentuated in autoML research, leading to a third limitation: a lack of ethical consideration—such as regarding algorithmic fairness—by all the included studies.

Fourth, inconsistent use of benchmarking represents a form of publication bias leading to erroneous conclusions of equivalent or superior autoML performance relative to conventional bespoke computational methods or clinicians. Many studies relied on historical controls or provided no benchmark at all. To confidently conclude that autoML performance compares well to bespoke models—and particularly to ‘state-of-the-art techniques’—a researcher with computational aptitude should have an opportunity to maximise performance. Finally, models should be deployed on separate datasets which were not used in testing or training, for external validation; this demonstrates generalisability, a critical component of clinical potential. Without external validation, overfitting to the datasets provided may lead to inflated estimates of performance (105). External validation is limited on many autoML platforms by a lack of ability to batch test on new data, or to export models for analysis and deployment.

### Limitations

This systematic review was limited by three issues: (**1**) PROBAST had to be adapted to apply it in non-diagnostic applications of autoML—we employed DECIDE-AI as a domain-specific quality indicator to mitigate this limitation, and utilised PROBAST in the context of trialling autoML technology rather than in validating models for clinical application. Development of more domain-specific tools to optimise AI-related systematic reviews is underway, and will be a welcome development (106,107).(**2**) Confidence in conclusions was affected by high risk of bias, a common theme in AI research more broadly (108). We provide comprehensive indicators of quality, risk of bias, and concerns regarding applicability to facilitate contextualisation of performance metrics. (**3**) It is difficult to draw conclusions for autoML as a modality because platforms are variable in their features, performances, and requirements—future reviews may focus on individual platforms, although the number of studies featuring most platforms is very small.

### Implications

Researchers applying a platform without providing benchmark comparators for the purposes of primary research or clinical work should justify their decision with validation data demonstrating that their approach is acceptable. Evidence should be contextually relevant, preferably pertaining to the same clinical task. While it is apparent that autoML has already begun to be applied in clinical research as a statistical tool, it is important that these tools are demonstrated to produce accurate, reliable, and fair models. Studies purported as evidence of validation of autoML are often limited by retrospective design, high risk of bias, and unfulfillment of conventional reporting standards—comparable to research regarding other AI technologies (109). Future comparative studies should address the limitations discussed above to convince researchers, clinicians, and policy makers that autoML platforms may be applied in lieu of bespoke modelling.

When reporting AI algorithms tasked with a certain clinical job, it would be helpful to avoid ambiguity in terminology. We would suggest a complete restriction of the terms ‘automated machine learning’, or ‘autoML’ for those algorithms built with technology that automates some or all parts of the process of the engineering process—all conventional ML models process data without human guidance, so description of these technologies as automated is redundant. Similar terms such as ‘automated artificial intelligence’, ‘automated machine learning’, and ‘automated deep learning’ are also redundant in the context of bespoke computational models. A simple alternative term for conventional ML projects would be ‘automatic’—these systems may automate a particular task, but their development is not automated, the defining feature of autoML.

The reduced barrier to entry in terms of computational expertise and hardware requirements conferred by many autoML platforms makes them a powerful contributor to democratisation of AI technology: a far greater number of clinicians and scientists are capable of ML development through use of these platforms. AutoML could be an invaluable resource for teaching, as individuals can more rapidly develop hands-on experience, learn by trial-and-error, and thereby develop intuitive understanding of the capabilities and limitations of ML. AutoML could also be applied in pilot studies, enabling clinicians with domain-specific expertise to explore possibilities for ML research—facilitating prioritisation of allocation of scarce resources such as GPU access and expert computer scientists. Validated platforms may be applied more broadly, including in patient care. Moreover, autoML is well placed to respond to calls to inculcate data-centric AI as opposed to model-centric development; focusing effort on curating high quality data, which limits development more often than code or model infrastructure (6). Acceleration in this process may be facilitated by large language models as their emerging capability to leverage plugins will allow autoML to facilitate AI building itself to fulfil user-defined aims (110).

Further work is indicated to improve validation of autoML platforms, either by allowing models to be exported, or by providing more comprehensive internal metrics. Other work should focus on improving the functionality of autoML, specifically on reducing the trade-offs currently implicit in selecting a platform with a given code intensity and computing locus. Using automation to reduce human error to optimise engineering and improve performance is one ideal: this has been demonstrated with structured data by AutoPrognosis. Increased functionality of code-free platforms while retaining the customisability of code-intense solutions is another ideal: H2O.ai Driverless AI offers the same functionality as the H2O.ai R and Python packages, but with a code-free graphical user interface. Alternatively, maximising accessibility by automating the whole engineering process may be desirable: Dedicaid is a platform requiring just data, with no customisable parameters, but has an ‘ethical compass’ which flags inappropriate datasets.

## Conclusion

AutoML performance is often comparable to bespoke ML and human performance. Many autoML platforms have been developed in academia and industry, with variable strengths and limitations. AutoML may prove especially useful in pilot studies and education, but potential use-cases include primary research and clinical deployment if platforms are rigorously validated. Future autoML research must be more transparently reported, adhere to reporting guidelines, and provide appropriate benchmarks for performance comparisons. Further autoML development should seek to minimise the ‘trade-offs’ currently inherent in selecting any given platform.

## Data availability statement

The raw data from this review may be provided upon request.

## Author contributions

AJT and DSWT conceived and coordinated the study. AJT, KE, LG, RH, YL, TFT, HC, and GL contributed to data collection. AJT and KE produced visualisations. AJT conducted data analysis. AJT and ZLT drafted the manuscript. KE, LG, RH, YL, TFT, GL, and DSWT edited the manuscript. All authors approved the final draft before submission.

