## Supplementary Material 1 for "Clinical performance of automated machine learning: a systematic review"

**Supplementary material 1—search strategy**

[1] = "autoML" OR "autoAI" OR "auto-ML" OR "auto-AI" OR "automated artificial intelligence" OR "automated machine learning" OR “auto-artificial intelligence” OR “auto-machine learning” OR “codeless” OR “code-free” OR “automated deep learning” OR “auto DL” OR “auto-DL” OR “automated neural network”

[2] = “medicine” OR “medical” OR “clinical” OR “healthcare” OR “diagnosis” or “diagnostic” OR “prognosis” OR “prognostic” OR "management" OR "treatment" OR "investigation"

*The Cochrane Library*: [1]

*Embase*: [1]

*MEDLINE*: [1]

*Scopus*: [1] AND [2]

Search conducted on 11 July 2022.
