## Supplementary Material 2 for "Clinical performance of automated machine learning: a systematic review"

| Inclusion | Exclusion |
|---|---|
| 1. Is published in the English language | 1. Is not published in the English language |
| 2. Is a peer-reviewed primary research article | 2. Is not a peer-reviewed primary research article<br>a. Review (systematic, scoping…)<br>b. Grey literature (e.g. preprint)<br>c. Conference paper<br>d. Commentary<br>e. Case report |
| 3. Is not a retracted article | 3. Is a retracted article<br>a. Misleading<br>b. Fraud<br>c. Unethical |
| 4. Utilises automated machine learning<br>a. Google, Microsoft, Apple, Sony, Amazon.<br>b. JADBio, H2O.ai.<br>c. Auto-WEKA, auto-Sklearn, autoKeras, TPOT, TuPaQ, ATM, Automatic Frankensteining, ML-Plan, Autostacker, AlphaD3M, Collaborative filtering<br>*List is not exhaustive! Use of any platform that automates any element of ML engineering is eligible.* | 4. Does not utilise automated machine learning<br>*No mention of autoML platform.* |
| 5. Algorithm is applied in a clinical context<br>a. Human patients<br>b. Related to disease diagnosis, management, or prognosis | 5. Algorithm is not applied in a clinical context<br>a. No aspect relevant to patients |

**Exclusion criteria are explicitly ranked**. Papers should be evaluated first by criterion 1, then 2, then 3 etc. When full-text screening, the first number ‘failed’ will need to be recorded. In Rayyan, this enables rapid exclusion by searching for e.g. ‘Italian’, ‘Japanese’ (non-English language), then ‘conference’, ‘comment’, ‘case report’ (non-peer reviewed research article).

**Examples are not exhaustive!** All published, English, peer-reviewed research articles testing any form of automated machine learning in a clinical context should be included. It may be useful to enter exclusion examples in the Rayyan search tool.
