## Supplementary figures and images for "Clinical performance of automated machine learning: a systematic review"

### Supplementary Material 5

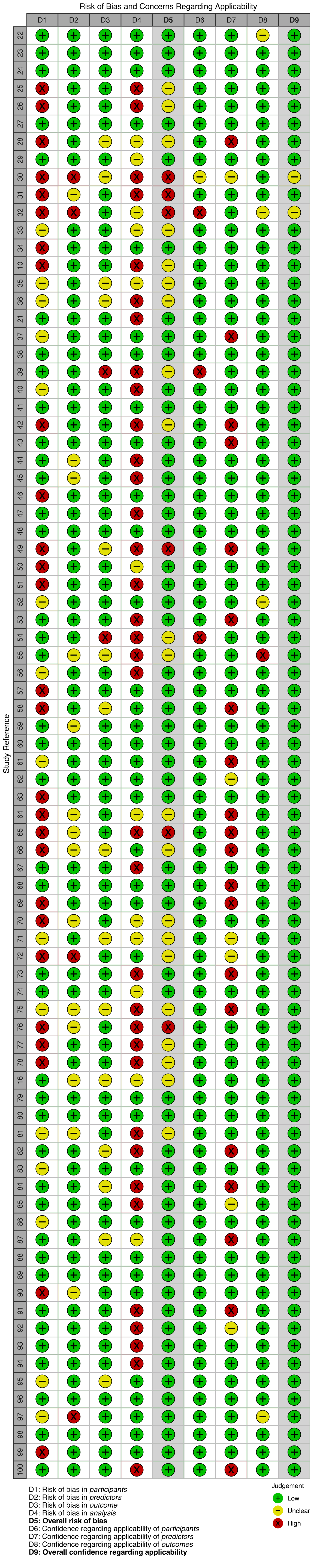
