## Supplementary Material 6 for "Clinical performance of automated machine learning: a systematic review"

| Platform | Specialty | Modality | Area under the precision-recall curve (95% CI) |
|---|---|---|---|
| Alaa and van der Schaar, 2018 | Pulmonology | | |
| AutoPrognosis | | AutoML | 0.58 (0.54 to 0.62) |
| TPOT | | AutoML | 0.51 (0.49 to 0.53) |
| SVM | | Bespoke computational | 0.50 (0.41 to 0.59) |
| Gradient boosting | | Bespoke computational | 0.55 (0.52 to 0.58) |
| Bagging | | Bespoke computational | 0.51 (0.47 to 0.55) |
| Nkam method | | Clinical | 0.50 (0.47 to 0.53) |
| Buzzetti method | | Clinical | 0.42 (0.40 to 0.44) |
| CF−ABLE−UK method | | Clinical | 0.28 (0.24 to 0.32) |
| FEV1% predicted criterion | | Clinical | 0.50 (0.48 to 0.52) |
| Antaki et al, 2021 | Ophthalmology | | |
| Cloud AutoML | | AutoML | 0.88 |
| Chen and Wujek, 2021 | Cardiology (1) | | |
| AutoDAL−SOAR | | AutoML | 0.81 |
| USDM | | AutoML | 0.71 |
| AER | | AutoML | 0.68 |
| Auto−WEKA | | AutoML | 0.65 |
| Auto−Sklearn | | AutoML | 0.77 |
| ASSL | | AutoML | 0.74 |
| | Cardiology (2) | | |
| AutoDAL−SOAR | | AutoML | 0.76 |
| USDM | | AutoML | 0.69 |
| AER | | AutoML | 0.71 |
| Auto−WEKA | | AutoML | 0.67 |
| auto−sklearn | | AutoML | 0.73 |
| ASSL | | AutoML | 0.71 |
| | Cardiology (3) | | |
| AutoDAL−SOAR | | AutoML | 0.79 |
| USDM | | AutoML | 0.76 |
| AER | | AutoML | 0.73 |
| Auto−WEKA | | AutoML | 0.69 |
| auto−sklearn | | AutoML | 0.73 |
| ASSL | | AutoML | 0.73 |
| | Cardiology (4) | | |
| AutoDAL−SOAR | | AutoML | 0.78 |
| USDM | | AutoML | 0.69 |
| AER | | AutoML | 0.72 |
| Auto−WEKA | | AutoML | 0.65 |
| auto−sklearn | | AutoML | 0.76 |
| ASSL | | AutoML | 0.73 |
| Danilatou et al, 2022 | Anesthesiology | | |
| JADBio | | AutoML | 0.74 |
| Faes et al, 2019 | Ophthalmology (1) | | |
| Cloud AutoML | | AutoML | 0.87 |
| | Ophthalmology (2) | | |
| Cloud AutoML | | AutoML | 0.99 |
| | Pulmonology (1) | | |
| Cloud AutoML | | AutoML | 1.00 |
| | Pulmonology (2) | | |
| Cloud AutoML | | AutoML | 0.57 |
| | Dermatology | | |
| Cloud AutoML | | AutoML | 0.93 |
| Ghosh et al, 2021 | Pulmonology | | |
| Cloud AutoML | | AutoML | 0.62 |
| Hu et al, 2022 | Hepatology (1) | | |
| TPOT | | AutoML | 0.76 |
| Radiomics pipeline | | Bespoke computational | 0.81 |
| | Hepatology (2) | | |
| TPOT | | AutoML | 0.77 |
| Radiomics pipeline | | Bespoke computational | 0.80 |
| Ikemura et al, 2021 | Pulmonology | | |
| H20.ai stacked ensemble | | AutoML | 0.81 |
| GBM | | Bespoke computational | 0.80 |
| DRF | | Bespoke computational | 0.78 |
| XGBoost | | Bespoke computational | 0.79 |
| XRT | | Bespoke computational | 0.78 |
| Kim et al, 2021 | Ophthalmology | | |
| Cloud AutoML 1 | | AutoML | 0.96 |
| Cloud AutoML 2 | | AutoML | 0.97 |
| Kumar et al, 2022 | Endocrinology | | |
| TPOT | | AutoML | 0.93 |
| Orlenko et al, 2020 | Cardiology (1) | | |
| TPOT | | AutoML | 0.88 |
| Logistic regression | | Bespoke computational | 0.83 |
| Decision tree | | Bespoke computational | 0.81 |
| Random forest | | Bespoke computational | 0.82 |
| | Cardiology (2) | | |
| TPOT | | AutoML | 0.78 |
| Logistic regression | | Bespoke computational | 0.73 |
| Decision tree | | Bespoke computational | 0.74 |
| Random forest | | Bespoke computational | 0.70 |
| Ou et al, 2021 | Neurology | | |
| TPOT | | AutoML | 0.63 (0.58 to 0.68) |
| Random forest | | Bespoke computational | 0.55 (0.46 to 0.63) |
| Logistic regression | | Bespoke computational | 0.43 (0.37 to 0.49) |
| ARSS | | Clinical | 0.50 (0.42 to 0.57) |
| Padmanabhan et al, 2019 | Cardiology (1) | | |
| Auto−Sklearn | | AutoML | 0.94 |
| Scikit learn | | Bespoke computational | 0.80 |
| | Cardiology (2) | | |
| Auto−Sklearn | | AutoML | 0.79 |
| Scikit learn | | Bespoke computational | 0.68 |
| Real et al, 2022 | Dentistry (1) | | |
| Auto−WEKA bagging | | AutoML | 0.86 |
| Auto−WEKA random committee | | AutoML | 0.90 |
| Auto−WEKA multilayer perceptron | | AutoML | 0.91 |
| | Dentistry (2) | | |
| Auto−WEKA logistic model tree | | AutoML | 0.92 |
| Auto−WEKA reduced error pruning tree | | AutoML | 0.79 |
| Auto−WEKA J48 | | AutoML | 0.75 |
| Auto−WEKA random tree | | AutoML | 0.89 |
| | Dentistry (3) | | |
| Auto−WEKA sequential minimal optimisation | | AutoML | 0.76 |
| Auto−WEKA multilayer perceptron | | AutoML | 0.74 |
| Auto−WEKA adaboost | | AutoML | 0.70 |
| Auto−WEKA bagging | | AutoML | 0.73 |
| Sakagianni et al, 2020 | Pulmonology | | |
| Cloud AutoML | | AutoML | 0.93 |
| Tahmasebi et al, 2021 | Breast | | |
| Cloud AutoML | | AutoML | 0.78 |
| Vagliano et al, 2022 | Anaesthesiology (1) | | |
| AutoPrognosis | | AutoML | 0.60 |
| Logistic Regression | | Bespoke computational | 0.58 |
| | Anaesthesiology (2) | | |
| AutoPrognosis | | AutoML | 0.60 |
| Logistic Regression | | Bespoke computational | 0.58 |
| APACHE | | Clinical | 0.52 |
| Wan et al, 2021 | Breast | | |
| Cloud AutoML | | AutoML | 0.95 |
| Random forest | | Bespoke computational | 0.90 |
| Convolutional neural network | | Bespoke computational | 0.88 |
| Logistic regression | | Bespoke computational | 0.75 |
| Linear discriminant analysis | | Bespoke computational | 0.66 |
| K-nearest neighbour | | Bespoke computational | 0.81 |
| Naïve Bayes | | Bespoke computational | 0.35 |
| SVM | | Bespoke computational | 0.67 |
| Adaboost | | Bespoke computational | 0.82 |
| Wang et al, 2020c | Otolaryngology | | |
| Cloud AutoML | | AutoML | 0.89 |
| Zeng and Zhang, 2020 | Breast | | |
| Cloud AutoML | | AutoML | 0.92 |

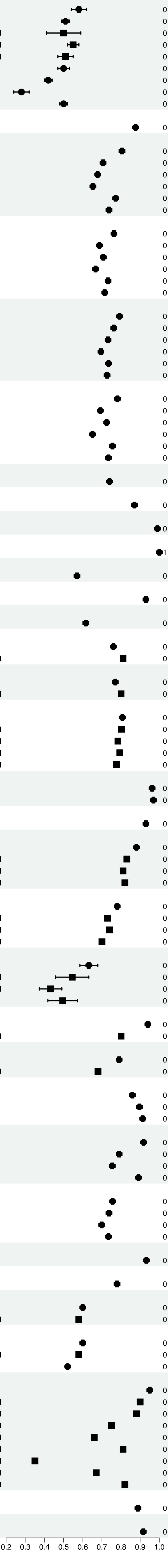


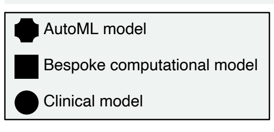
